# The effectiveness of a FY-1 doctor preparation course for final year medical students

**DOI:** 10.1101/2020.08.24.20180109

**Authors:** William Beedham, Kasun Wanigasooriya, Georgia R. Layton, Ley Taing Chan, Adnan Darr, Devender Mittapalli

## Abstract

**Background:** Starting work as a junior doctor can be daunting for any medical student. There are numerous aspects of the hidden or informal curriculum which many students fail to acquire during their training.

**Objectives:** To evaluate the effectiveness of a novel foundation year one (FY1) doctor preparation course focusing on certain core topics, practical tips and components of the hidden curriculum. The primary objective was to improve the confidence level and knowledge of final year medical student transition to FY1 doctors.

**Method:** A two-day, practical course titled “Preparation 2 Practice” delivering hands-on, small-group and lecture-based teaching, covering core medical student undergraduate curriculum topics in medicine and surgery. The course content spanned therapeutics, documentation skills and managing acute clinical tasks encountered by FY1 doctors during an on-call shift. A pre- and post-course survey and knowledge assessment were carried out to assess the effectiveness of the course. The assessment was MCQ-based, derived from topics covered within our course. The 20-question test and a short survey were administered electronically.

**Results:** Twenty students from a single UK medical school attended the course. 100% participation was observed in the pre- and post-course test and survey. The median post-course test result was 22 (IQR 20.25 –23.75) which was higher than the median pre course test score of 18.75 (IQR 17–21.75). A Wilcoxon sign rank test revealed a statistically significant difference between the pre- and post-course test results (p=0.0003). The self-reported confidence score of delegates on starting work as a junior doctor was also significantly higher following the course (p=0.004).

**Conclusion:** The results show a significant improvement in perceived confidence and knowledge on core curriculum topics amongst final year medical students having attended our FY1 doctor preparation course. We conclude that there is scope for similar supplementary courses as an adjunct to the undergraduate medical curriculum.

## Introduction

The transition from medical student to foundation year one (FY1) doctor can be daunting and challenging. Despite 5 to 6 years of undergraduate medical training comprising of countless hours learning theory and hands-on practical teaching in the clinical environment, this transition is still associated with a steep learning curve. This may in part be attributed to a lack of preparation for this transition (Monrouxe 2018). The General Medical Council (GMC), the regulator of doctors in the UK, reports in their 2019 guidance titled “The state of medical education and practice in the UK”, that a lack of preparedness for FY1 is associated with long term effects such as doctor burnout (Anwar et al. 2019). Consequently, the GMC has placed significant emphasis on the need for “transition interventions” to maximise medical student preparedness before starting FY 1 jobs.

A lack of preparation for some of the vocational aspects demanded by the FY1 job, particularly within components of the hidden curriculum (Cameron et al. 2014) of medical student training, is frequently detected amongst newly qualified doctors. Lempp et al. define the hidden curriculum as, “processes, pressures and constraints which fall outside the formal curriculum, and which are often unarticulated or unexplored” (Lempp and Seale 2003). Lack of exposure to and training in components within the hidden curriculum may result in FY1 doctors perceiving themselves as “unprepared” (Kellet et al. 2015) which can often be viewed by senior clinicians as incompetence (Matheson and Matheson 2009). These perceptions in combination with a noticeable but short rise in hospital-based mortality in the first few weeks of August, as new FY1 doctors take to their roles as doctors for the first time, have contributed to the origin of the term “Black-Wednesday” (Gaskell et al. 2016) in the UK. This poor preparation not only has an impact on patient care but also impacts the individual doctors and their teams at a personal and professional level (Burridge et al. 2020). Sadly, there is no way of completely removing the responsibility that new doctors are about to assume. However, improving preparedness for this transition may enhance both patient and doctor experience (Yardley et al. 2018).

Equipping final year medical students with hospital-specific knowledge which covers areas on the “informal curriculum” of medicine - typically not covered by medical schools - should allow students to feel more confident and prepared to transition into the junior doctor role (Ozolins et al. 2008). This concept of providing senior medical students with vocational training before they start their foundation jobs is not novel, but is far from common practice (Monrouxe et al. 2017). The introduction of a one-week or longer shadowing programme prior to commencing work as a FY1, often termed an assistantship, is one such example. Courses and e-learning tools exist which cover practical elements and common principles relating to the management of acutely unwell patients (Cullinan et al. 2017). Several have also focused on a more simulation-based style of teaching to ease this transition (Teagle et al. 2017; Morgan et al. 2017). An increasing demand on healthcare services and fears of litigation have resulted in a significant reduction in the hands-on vocational training opportunities for medical students as well as junior doctors (Roberts 2004). Numerous courses (Newby et al. 2019) and simulation training sessions (Watmough et al. 2016) have been created to fill these gaps in learning at various stages of medical training. Even though these courses are not a substitute to formal undergraduate or postgraduate medical training, there is a growing demand for them to reinforce and supplement medical training at every stage (Humm et al. 2018). Therefore, a course for final year medical students which ensures the revision of key topics relating to the management acutely unwell patients and other components of the hidden curriculum could act as a supplement to aid this difficult transition. Therefore, a course designed for final year medical students which ensures the revision of key topics relating to the management acutely unwell patients and other components of the hidden curriculum could act as a supplement to aid this difficult transition.

Our course, “The Master Surgeon Preparation 2 Practice Course” (P2P) was developed to cover a range of select concepts which had been identified from published literature as poorly understood by previous FY1 doctors (Goldacre et al. 2010). Furthermore, the results from an initial survey of an electronic on-call job list for junior doctors at an acute NHS Hospital Trust in the UK, which was conducted by the authors was used as a guide to design the course curriculum. The course programme comprised of topics based on the most commonly encountered tasks requested of a FY1 doctor whilst on-call, as identified during the former survey. **Figure-1** highlights the topics covered within this course. This internal quality control study aims to assess the effectiveness of this course at improving student knowledge and confidence immediately prior to starting work as a FY1 doctor.

**Figure 1:**
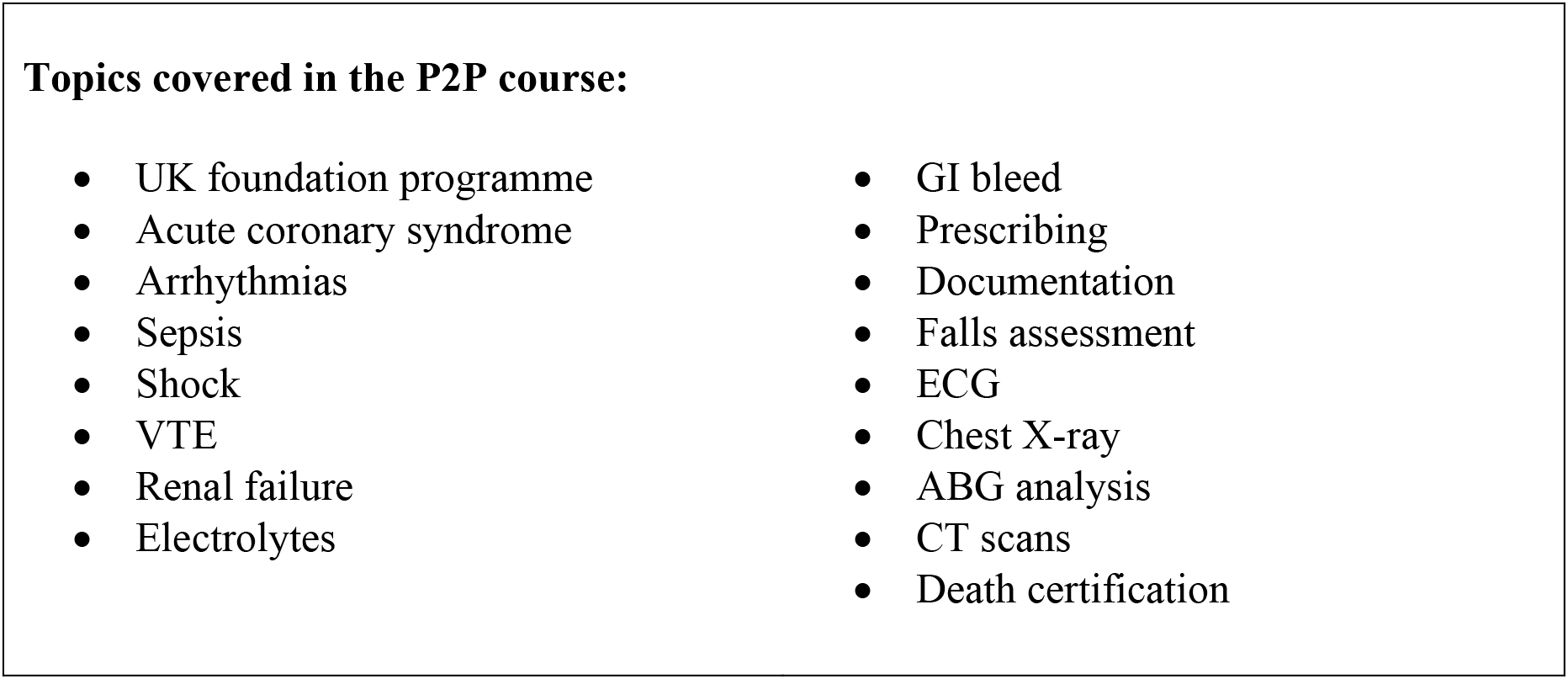
Topics covered in the P2P course.

## Methods

The two-day P2P course was run for final year medical students attending a single medical school in the UK. The course was advertised through social media platforms, email and word of mouth. Twenty places were offered to students and was delivered free of charge to attendees. The course content was delivered by a faculty comprising of doctors of all grades from FY1 to consultants currently employed within the NHS. Course content was delivered in the form of lectures, small group tutorials, practical stations and quizzes. Care was taken for each session to be delivered by a mix of one junior and one senior doctor to amalgamate peer and expert teaching. All course content was peer-reviewed and validated by at least two independent reviewers prior to delivery. The course programme can be found in **Appendix 1**.

Pre- and post-course tests were administered to participants electronically using SurveyMonkey™ (San Mateo, California, USA). The questions within these two tests were kept identical with the aim of accurately assessing a change in knowledge and confidence of participants. The peer-reviewed test comprised of multiple-choice questions (MCQ), assessing the knowledge from key surgical and medical topics covered within this course. Alongside the MCQ tests, participants were also given a 5-point Likert scale survey to assess confidence levels before and after the course, ranging from not very confident at all (1), to very confident (5).

Statistical analysis was performed using SPSS (IBM, Armonk, New York, USA) and Microsoft Excel (Redmond, Washington, USA). The null hypotheses were as follows; 1) there was no significant difference between pre- and post-course test scores, taken before and after the P2P course respectfully and 2) no significant difference between pre- and post-course participant perceived confidence level of starting work as a FY1 doctor. A p-value of <0.05 was assigned as the threshold for statistical significance.

## Results

Twenty final year medical students completed our two-day course. All twenty students also completed the pre- and post-course tests and surveys. The students were all at the end of their medical training and were awaiting graduation. The results of the pre- and post-course tests have been demonstrated in **Table 1 and Figure 2**. The median participant test score increased from 18 (IQR = 17-21.75) to 22 (IQR = 20.25-23.75). A Wilcoxon sign rank test demonstrated that there was a statistically significant difference in the pre and post-course test scores (Z=-3.646, p = 0.0003). The course led to an increase in participant knowledge in the key topics covered. The results for the pre- and post-course confidence level survey can be seen in **Table 1**. The median pre- and post-course confidence scores were 3 (IQR 3-2) and 4 (IQR 4-3) respectively. A Wilcoxon sign rank test again demonstrated that there was a statistically significant difference between pre- to post-confidence scores of participants (Z = −3.578, p = 0.004), demonstrating an increase in participant confidence.

**Table 1:** Pre and post-course test and survey scores. Demonstrates a significant increase in test score and confidence after the course.

| **Participant test scores** |  |  |
| --- | --- | --- |
|  | **Pre-course** | **Post-course** |
| **Median** | 18 | 22 |
| **Q1** | 17 | 20 |
| **Q3** | 21 | 23 |
| **IQR** | 21-17 | 23-20 |
| **Std. Deviation** | 2.62 | 2.33 |

| **Self-reported confidence of starting work (1-5 Likert Scale)** |  |  |
| --- | --- | --- |
|  | **Pre-course** | **Post-course** |
| **Median** | 3 | 4 |
| **Q1** | 2 | 3 |
| **Q3** | 3 | 4 |
| **IQR** | 3-2 | 4-3 |
| **Std. Deviation** | 0.733 | 0.671 |

**Figure 2:**
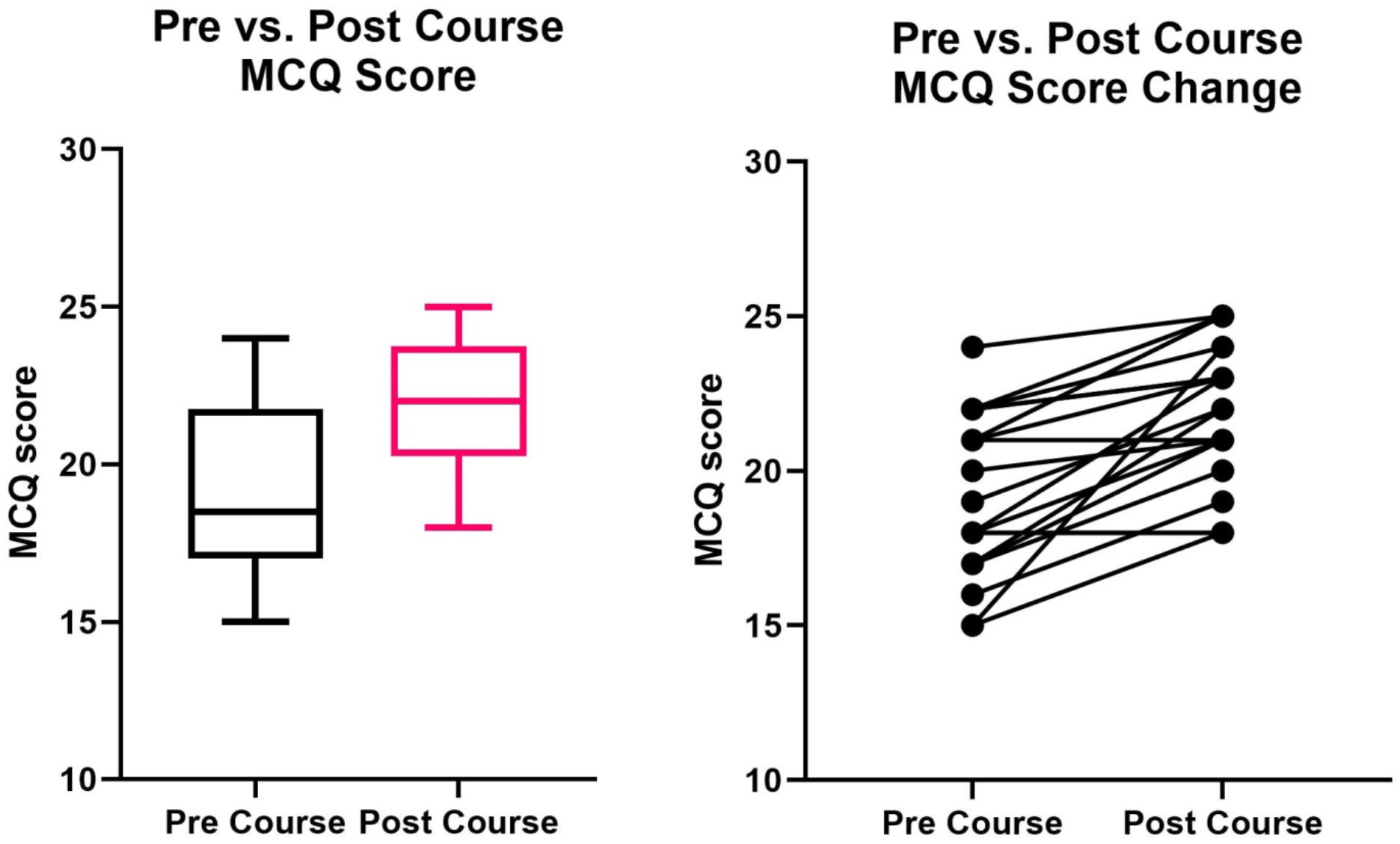
Student pre- and post-course MCQ scores. Demonstrates significant improvement in pre- to post-course test scores.

## Discussion

Several authors have identified the positive outcomes of transition intervention courses for final year medical students starting work as new junior doctors. Blencowe et al. demonstrated that 97% of FY1 doctors who attended their comparable “targeted structured induction programme” felt prepared for practising medicine afterwards (Blencowe et al. 2015). Importantly, they followed this cohort throughout their initial employment and identified that the incidence of errors reported by the FY1s who attended their course fell by 45% in the first 4 months of their jobs compared to those that did not. Teagle et al. also demonstrated a significant increase in prescribing confidence in students attending their preparation for practice course (Teagle et al. 2017).

Our findings in line with previously published literature would suggest that this course, or a suitable alternative could have a role to play as an adjunct to the current medical student curriculum at this particular University. It was clear from our results that this course went some way to address short comings of the current curriculum. Following the two-day course, participants scored significantly higher on a knowledge test and rated their self-perceived confidence as significantly higher. We can therefore hypothesise, by extrapolation, that participants should demonstrate greater knowledge of key medical and surgical topics that were covered during the course when starting as FY1 doctors.

Although the current undergraduate medical student curricula across the UK include some form of “transition intervention” such as the assistantship, which provides students with a degree of vocational exposure, we believe that this simple exposure is not sufficient. A more structured approach to teaching these practical elements is required. These courses and ours alike promote a collaborative approach to learning by utilising group interaction, discussion and the opportunity for sharing of anecdotal experience. The use of recent graduates, as well as senior doctors, to deliver the course content capitalises on the benefits of near-peer teaching, a methodology proven to be effective (Gottlieb et al. 2016).

The essential topics covered in this course have often failed to garner significant attention within the formal undergraduate curriculum for medical students. Without focused teaching through courses such as ours, acquisition of this essential knowledge is inevitably postponed until FY1s are exposed in the real-world, to the clinical scenarios taught during this course. Such encounters can be challenging, stressful and could impact patient care, and lead to adverse effects on new doctor wellbeing. Therefore, the purpose of these courses is to address such concerns within a safe environment before medical students commence work as FY1s. Furthermore, they ensure that students have fewer knowledge deficits and demonstrate greater confidence at starting their new jobs. Hence, such courses also complement the GMC’s call for a smooth transition from final year medical student to FY1 (Anwar et al. 2019).

To our knowledge, our course was the first of its kind designed for undergraduates in the West Midlands. The greatest strength of this course was its breadth and succinctness. Compared to other courses which cover specific topics such as prescribing or acute medicine our P2P course covered a range of topics, essentially making it a “crash course” for the new FY1. Other courses are also run over several days to weeks. We acknowledge that time before starting FY 1 is often viewed as a final period of leisure before initiating a lifelong career as a doctor. Therefore, a shorter and more concise course was deemed likely to be better received.

The limitations of our study included a small sample size. This carries both the disadvantage of reduced knowledge dissemination and lower accuracy of any statistically significant findings from our analysis. However, it did allow for a high ratio of faculty to students which better supported a personalised learning experience and more opportunities for discussion. The latter may also explain the observed results and its perceived effectiveness. The pre- and post-course assessments were not administered under exam conditions, potentially reducing the reliability of the participant test scores. Whilst the use of identical pre- and post-course questions ensured standardisation in testing, this is a limited approach, permitting participants to consider the answers to questions during the delivery of the session. Our course did not offer clinical skills training or simulation-based training. However, students were offered numerous small group practical stations (e.g. prescribing, documentation, certifying death) within the course where individual students were given the opportunity to practice performing these tasks using case-based scenarios using prescription charts, history sheets and death certificates.

With such courses, there is a significant risk of selection and responder bias. The attendees of this course, by nature, are more likely to be conscientious students who may be above average performers at baseline. These results are therefore not generalisable to all medical students. Our cohort of participants were not followed over a period of time. Therefore, it was not possible to formally assess whether the attendance at this course led to a sustained increase in knowledge and confidence. This is a key area for further research to objectively assess the long-term impact of transition interventions such as these preparation for practice courses.

## Conclusion

Our findings combined with those of the GMC, make it clear that many final year medical students feel unprepared to start work as a FY1 doctor. Whether perceived or real, such beliefs can result in junior doctors not achieving their full potential and negatively impact their future practice. Furthermore, if this lack of preparation and confidence translates to an inability to perform effectively within the FY1 role, then patientcare may be compromised. Having identified this knowledge gap within the undergraduate medical school curriculum we designed and delivered a course which a group of final year medical students found to be extremely useful. Our data demonstrated that this course was effective at increasing attendee knowledge and at improving their confidence in starting work as FY1s.

Additional research comprising of larger sample sizes, randomised control trials and longer-term cause-effect outcome analysis is required before robust recommendations can be made regarding the integration of this type of course into the formal undergraduate curriculum. Until then, these types of independent courses and training programmes are likely to play a significant role in helping medical students improve their confidence and knowledge of key vocational topics towards the end of their undergraduate medical training.

### Take Home Messages

- The transition between medical student is daunting with many students feeling unprepared.
- There is a hidden curriculum in medicine, which is often missed by students.
- Our course addressed areas of the hidden curriculum, which improved student knowledge and made them feel better prepared for becoming a doctor.
- A course such as ours should be included in the medical undergraduate curriculum.

## Data Availability

All relevant data has been included within the tables of the manuscript. A full data set is in the possession of the authors.

## Ethical approval

This project was conducted as part of an internal quality control audit of The Master Surgeon Trust (UK Charity EW0332), which developed, organised and ran this course. The audit process was authorised by the charity executive committee. The Medical Research Council, Health Research Authority “Is my study research?” online tool (http://www.hra-decisiontools.org.uk/research/) was used to confirm that this study was not research. Therefore, research ethics committee review or health research authority approval was not required.

## Notes on contributors

W Beedham: wrote the manuscript, and interpreted data.

K Wanigasooriya: designed and ran course, data collection, statistical analysis, editing manuscript

GR Layton: Editing manuscript, data interpretation.

A Darr: Designed and ran course, editing manuscript

LT Chan: Designed and ran course, data collection.

D Mittapalli: senior supervision, data interpretation, editing manuscript.

